# Detection of SARS-CoV-2 lineage P.1 in patients from a region with exponentially increasing hospitalization rates in February 2021, Rio Grande do Sul, Southern Brazil

**DOI:** 10.1101/2021.03.09.21253204

**Authors:** Andreza Francisco Martins, Alexandre Prehn Zavascki, Priscila Lamb Wink, Fabiana Caroline Zempulski Volpato, Francielle Liz Monteiro, Clévia Rosset, Fernanda De-Paris, Álvaro Krüger Ramos, Afonso Luís Barth

## Abstract

The emergence SARS-CoV-2 P.1 lineage has been coincidental with a rapid growth in hospitalization in the northern region of Brazil. An exponential growth of severe COVID-19 occurred in Rio Grande do Sul state, Southern Brazil in February-2021. Whole-genome sequencing revealed that the previously undetected P.1 lineage accounted for 88.9% of specimens collected from patients at a referral COVID-19 hospital. These findings raise concerns regarding a possible association between lineage P.1 and rapid growth in cases and hospitalizations.

---

The state of Amazonas, Northern Brazil, has been hit hardest by COVID-19 among all regions in the country. The first wave was mostly driven by lineage B.1.195, which was subsequently replaced by B.1.1.28 [1]. A second wave with striking exponential growth in hospitalizations and deaths has been observed since mid-December 2020, and has been shown to coincide with the emergence of the P.1 variant of concern (VOC), which was first detected in a Brazilian traveler from Amazonas to Japan on January 2, 2021 [2]. Rio Grande do Sul (RS) is the southernmost Brazilian state. With a population of 11,422,973 and land borders with Uruguay and Argentina, it was one of the least affected states during the first wave (early July–September 2020), although the toll increased markedly from early November to late December 2020 [3]. A previous study conducted in the municipality of Esteio, RS, from May to October 2020 revealed that the most common lineages were B.1.1.33 and B.1.1.248 [4]. Genomic surveillance by the central state laboratory confirmed the predominance of these lineages across RS, following a distribution similar to that observed in other Brazilian states, but revealed that lineage P.2, first detected in October 2020, had been gradually replacing these as the predominant variant as of 31 January 2021 [5]. Lineage P.1 was first detected in a single clinical specimen (of 23 collected in the year 2021 to date) obtained on 01 February 2021 from a resident of Gramado, a popular tourist destination in RS, who however had no history of travel or contact with returning travelers from Northern Brazil [6].

Since epidemiological week 6, starting 07 February 2021, RS has witnessed exponential growth of hospitalizations for COVID-19, from 1738 inpatients on 07 February to 6995 on 06 March—a 3.8-fold increase in the number of hospitalized patients with COVID-19 (Figure 1), resulting in collapse of the regional health system. Since community transmission of lineage P.1 was documented in a highly visited RS municipality one week before the exponential increase in hospitalizations (Figure 1), we decided to investigate whether the P.1 VOC could be detected in specimens from patients hospitalized on February 2021 and compare our findings with specimens obtained in January 2021 and the previous year from a large tertiary care center in the state capital, Porto Alegre, which has been designated as a COVID-19 referral center.

**Figure 1.**
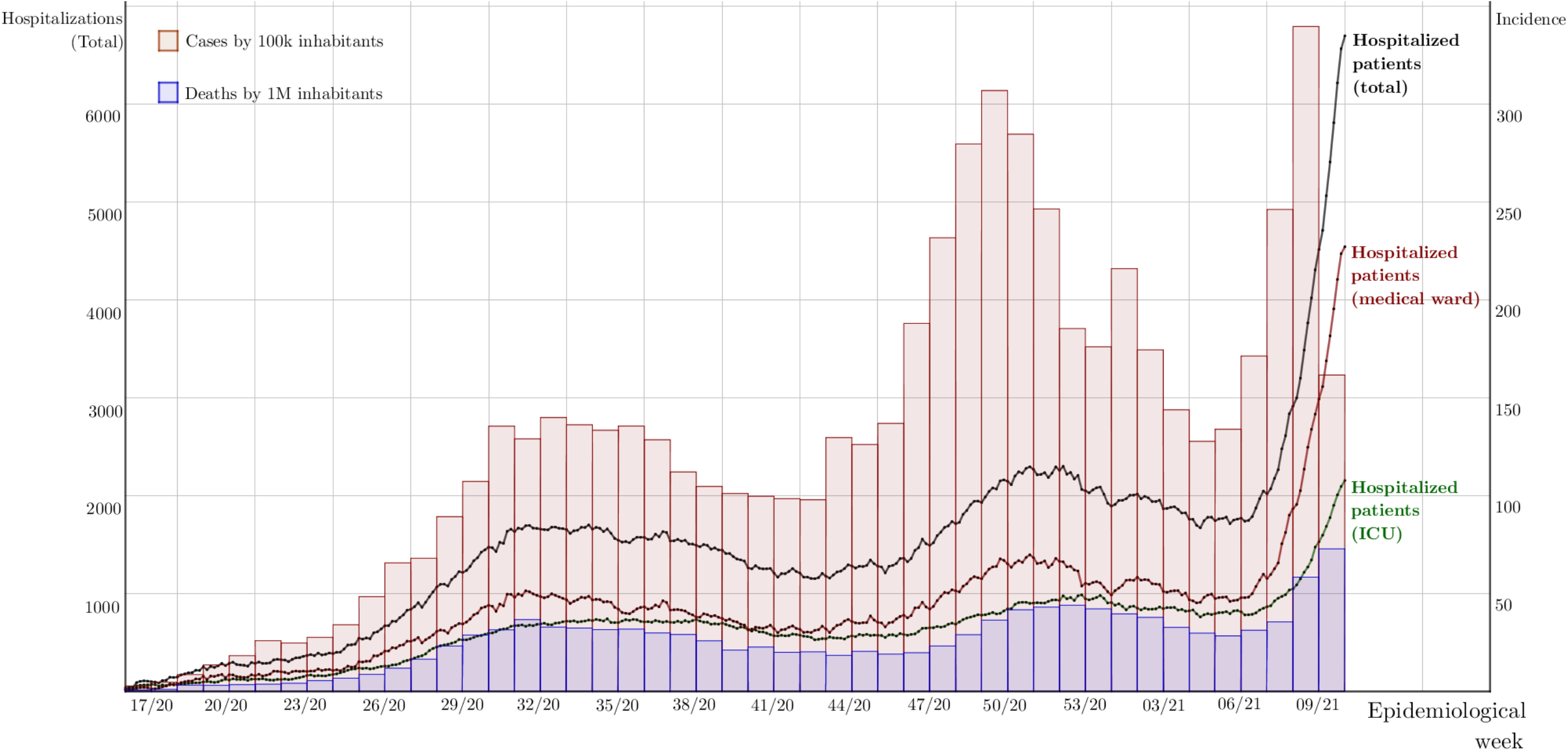
Number of hospitalized patients, cases, and deaths in Rio Grande do Sul, April 12, 2020–present. From epidemiological week 06-2021 (February 7-16, 2021) through week 09-2021 (February 28-March 6, 2021), the total number of hospitalized patients with a positive diagnostic test for COVID-19 exhibited exponential growth, with an average doubling time of 13.4 days and a daily growth rate of 5.29%. In the same period, similarly exponential growth (doubling time 13.3 days, daily growth rate 5.35%) was observed for hospitalizations in the city of Porto Alegre, where most P.1 specimens were found (not shown in figure). Note that cases and deaths for epidemiological weeks 08-2021 and 09-2021 are not final and are subject to increase due to the lag in reporting.

Of 337 specimens obtained from hospitalized patients or hospital workers in February 2021 and 307 obtained in January 2021, 27 (8.0%) and 15 (4.9%), respectively, were selected for genomic analysis based on cycle threshold (Ct) values (lower values were selected) of the RT- qPCR (https://www.fda.gov/media/134922/download) and patient age (specimens from younger patients were preferred). Of 3224 specimens obtained in 2020, 26 (0.81%) were randomly selected among those with lower Ct values.

Sequencing libraries were prepared with the CleanPlex SARS-CoV-2 panel (Paragon Genomics, Inc, Hayward, CA, USA) protocol for target enrichment and library preparation, following manufacturer instructions (https://www.paragongenomics.com/wp-content/uploads/2020/03/UG4001-01_-CleanPlex-SARS-CoV-2-Panel-User-Guide.pdf). The resulting libraries were pooled in equimolar amounts and were 250bp-paired-end read-sequenced in an Illumina MiSeq sequencer (Illumina, San Diego, CA, USA). Consensus sequences were generated by the QIASeq SARS-CoV-2 pipeline (QIAGEN CLC Genomics Workbench 21). We obtained 68 whole genomes with high quality (coverage >80%, <10% N, >29.5 Kb). The lineages were classified using *pangolin* (accessed on 28-02-2021) [7]. The sequences were deposited into GISAID database (https://www.gisaid.org/)

Of 27 specimens obtained in February 2021, 24 (88.9%) were identified as lineage P.1, versus one (6.7%) of 15 specimens obtained in January 2021. Data regarding the P.1 specimens are listed in Table 1. Thirteen patients (46.4%) were 20 to 29 years old, five (17.8%) were 30-39, four (14.3%) were 50-59, two (7.1%) were 40-49, two (7.1%) were 60-69, and two (7.1%) were children under 2.

**Table 1.**
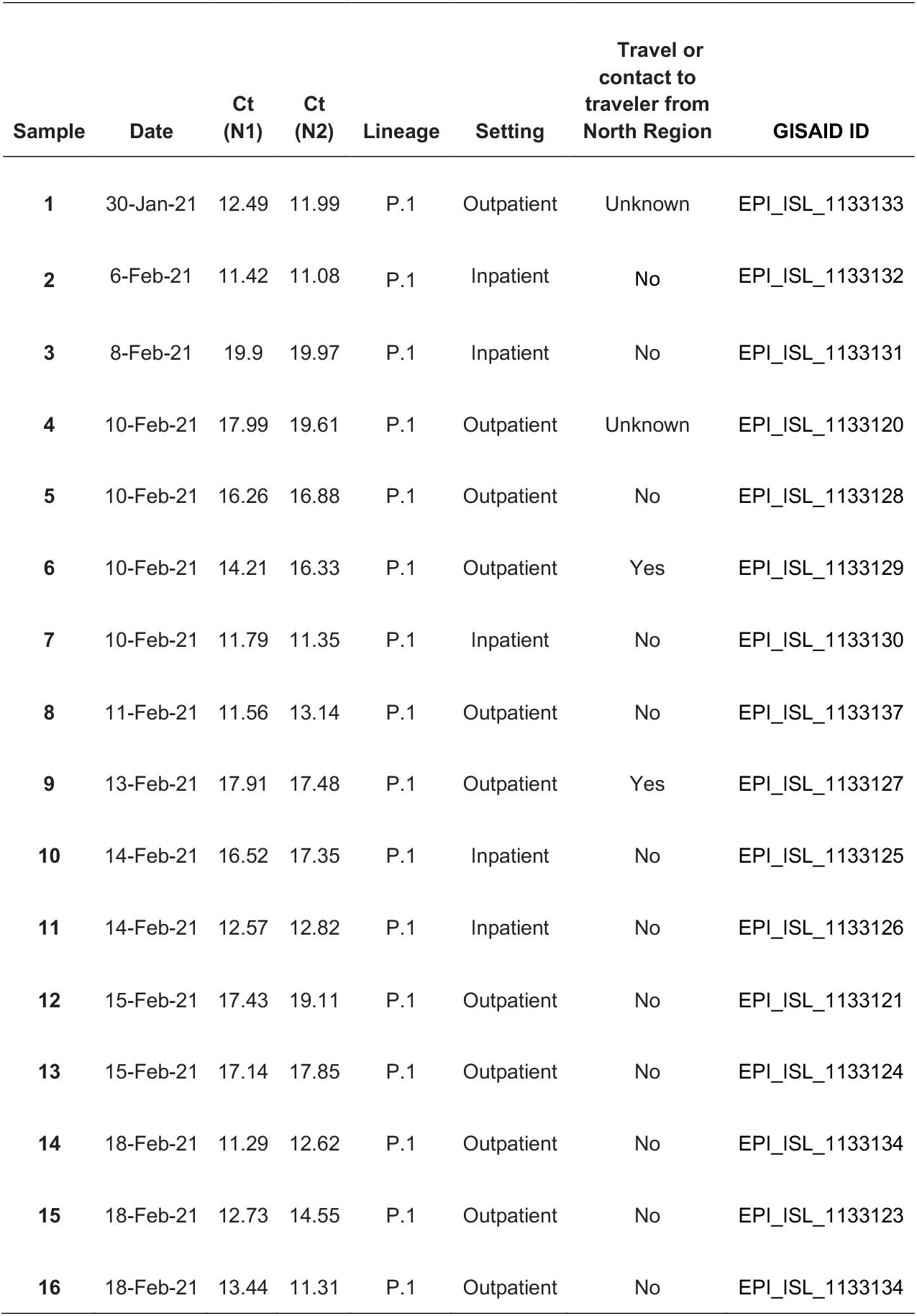
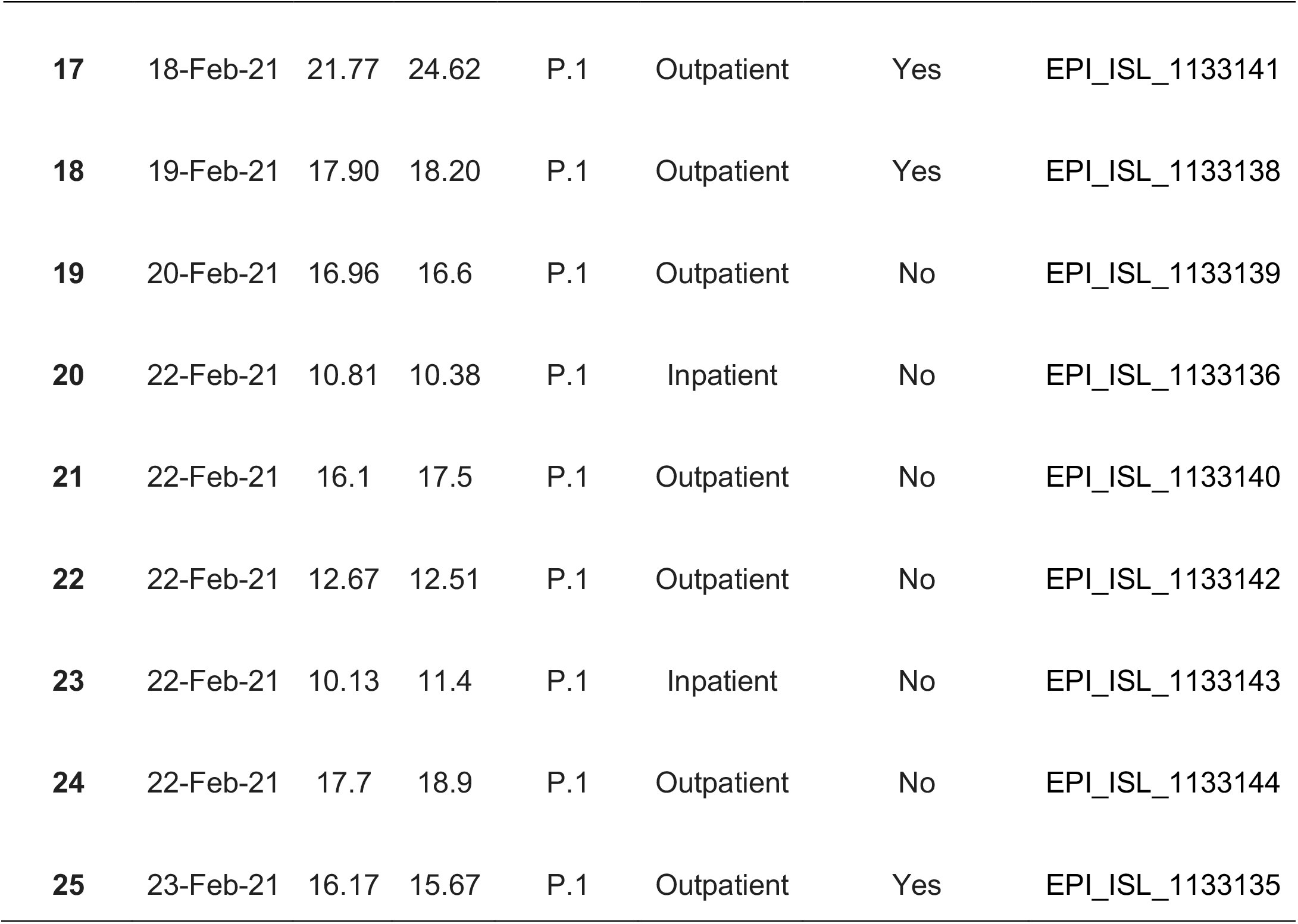
Characteristics of P.1 specimens, setting of specimen recovery, and epidemiological history of patients.

| Sample | Date | Ct (N1) | Ct (N2) | Lineage | Setting | Travel or contact to traveler from North Region | GISAID ID |
| --- | --- | --- | --- | --- | --- | --- | --- |
| 1 | 30-Jan-21 | 12.49 | 11.99 | P.1 | Outpatient | Unknown | EPI_ISL_1133133 |
| 2 | 6-Feb-21 | 11.42 | 11.08 | P.1 | Inpatient | No | EPI_ISL_1133132 |
| 3 | 8-Feb-21 | 19.9 | 19.97 | P.1 | Inpatient | No | EPI_ISL_1133131 |
| 4 | 10-Feb-21 | 17.99 | 19.61 | P.1 | Outpatient | Unknown | EPI_ISL_1133120 |
| 5 | 10-Feb-21 | 16.26 | 16.88 | P.1 | Outpatient | No | EPI_ISL_1133128 |
| 6 | 10-Feb-21 | 14.21 | 16.33 | P.1 | Outpatient | Yes | EPI_ISL_1133129 |
| 7 | 10-Feb-21 | 11.79 | 11.35 | P.1 | Inpatient | No | EPI_ISL_1133130 |
| 8 | 11-Feb-21 | 11.56 | 13.14 | P.1 | Outpatient | No | EPI_ISL_1133137 |
| 9 | 13-Feb-21 | 17.91 | 17.48 | P.1 | Outpatient | Yes | EPI_ISL_1133127 |
| 10 | 14-Feb-21 | 16.52 | 17.35 | P.1 | Inpatient | No | EPI_ISL_1133125 |
| 11 | 14-Feb-21 | 12.57 | 12.82 | P.1 | Inpatient | No | EPI_ISL_1133126 |
| 12 | 15-Feb-21 | 17.43 | 19.11 | P.1 | Outpatient | No | EPI_ISL_1133121 |
| 13 | 15-Feb-21 | 17.14 | 17.85 | P.1 | Outpatient | No | EPI_ISL_1133124 |
| 14 | 18-Feb-21 | 11.29 | 12.62 | P.1 | Outpatient | No | EPI_ISL_1133134 |
| 15 | 18-Feb-21 | 12.73 | 14.55 | P.1 | Outpatient | No | EPI_ISL_1133123 |
| 16 | 18-Feb-21 | 13.44 | 11.31 | P.1 | Outpatient | No | EPI_ISL_1133134 |
| **17** | 18-Feb-21 | 21.77 | 24.62 | P.1 | Outpatient | Yes | EPI_ISL_1133141 |
| **18** | 19-Feb-21 | 17.90 | 18.20 | P.1 | Outpatient | Yes | EPI_ISL_1133138 |
| **19** | 20-Feb-21 | 16.96 | 16.6 | P.1 | Outpatient | No | EPI_ISL_1133139 |
| **20** | 22-Feb-21 | 10.81 | 10.38 | P.1 | Inpatient | No | EPI_ISL_1133136 |
| **21** | 22-Feb-21 | 16.1 | 17.5 | P.1 | Outpatient | No | EPI_ISL_1133140 |
| **22** | 22-Feb-21 | 12.67 | 12.51 | P.1 | Outpatient | No | EPI_ISL_1133142 |
| **23** | 22-Feb-21 | 10.13 | 11.4 | P.1 | Inpatient | No | EPI_ISL_1133143 |
| **24** | 22-Feb-21 | 17.7 | 18.9 | P.1 | Outpatient | No | EPI_ISL_1133144 |
| **25** | 23-Feb-21 | 16.17 | 15.67 | P.1 | Outpatient | Yes | EPI_ISL_1133135 |

The other three specimens from February 2021 were identified as lineages B.1.1.28 (n=2) and B.1.1.143 (n=1). The lineages identified in January and February 2021 and throughout 2020 are listed in Table 2. Among the 68 sequenced specimens, 43 (63.2%) and 25 (35.8%) were from inpatients and outpatients, respectively; there was no significant difference in proportion of the specimen recovery setting across the periods of study (P=0.46; chi-square test).

**Table 2.**
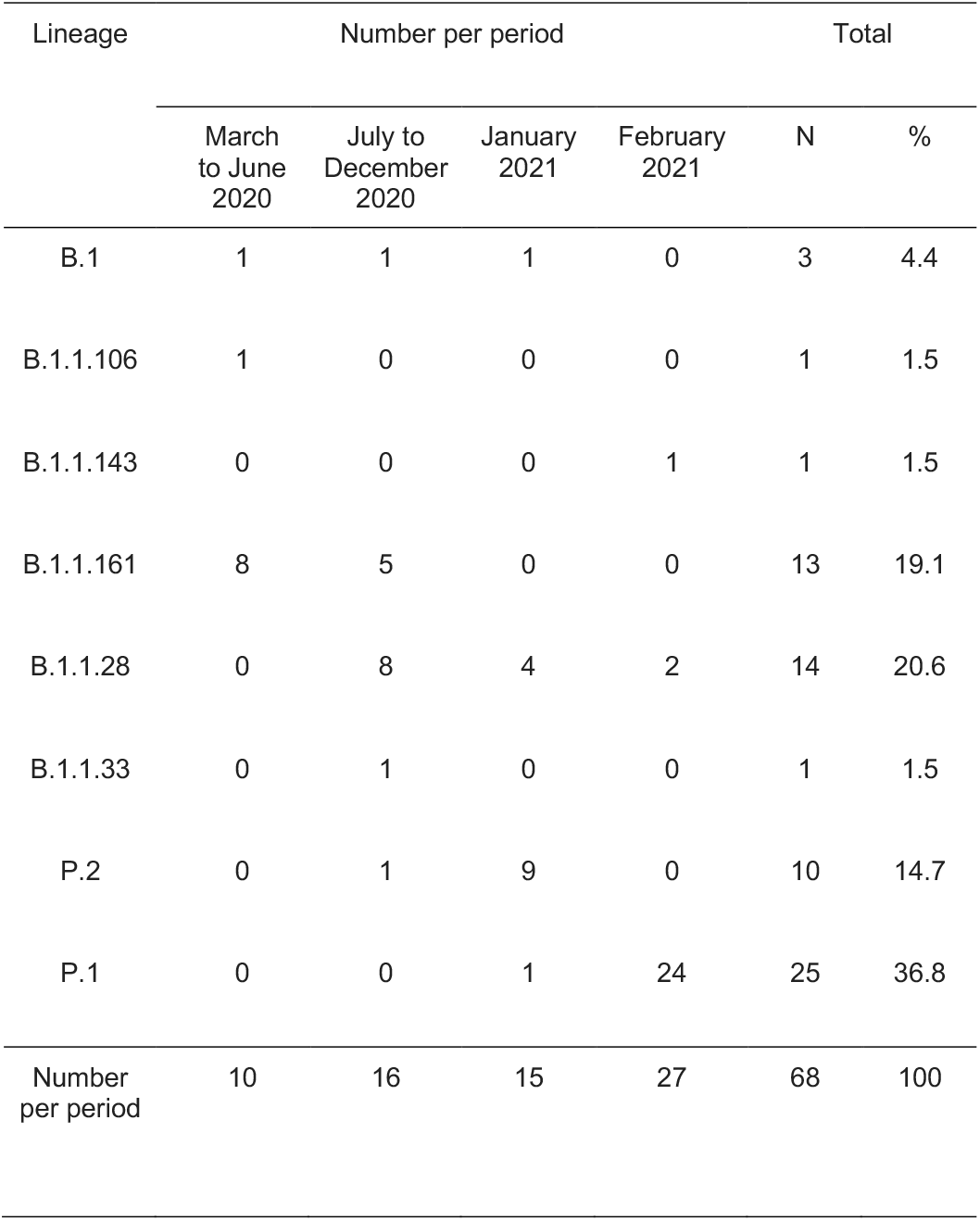
Distribution of lineages across distinct periods of the study.

| Lineage | Number per period |  |  |  | Total |  |
| --- | --- | --- | --- | --- | --- | --- |
|  | March to June 2020 | July to December 2020 | January 2021 | February 2021 | N | % |
| B.1 | 1 | 1 | 1 | 0 | 3 | 4.4 |
| B.1.1.106 | 1 | 0 | 0 | 0 | 1 | 1.5 |
| B.1.1.143 | 0 | 0 | 0 | 1 | 1 | 1.5 |
| B.1.1.161 | 8 | 5 | 0 | 0 | 13 | 19.1 |
| B.1.1.28 | 0 | 8 | 4 | 2 | 14 | 20.6 |
| B.1.1.33 | 0 | 1 | 0 | 0 | 1 | 1.5 |
| P.2 | 0 | 1 | 9 | 0 | 10 | 14.7 |
| P.1 | 0 | 0 | 1 | 24 | 25 | 36.8 |
| Number per period | 10 | 16 | 15 | 27 | 68 | 100 |

As a post-hoc analysis, we also compared the Ct values of the N1 and N2 targets across the 12-month period of the study. Ct values in February 2021 were the lowest observed in the period (mean, 21.11; 95%CI, 20.39-21.84; P=0.14; and 22.01; 95%CI, 21.23-22.78; P<0.001, for N1 and N2 targets, respectively; one-way ANOVA; Figure 2).

**Figure 2.**
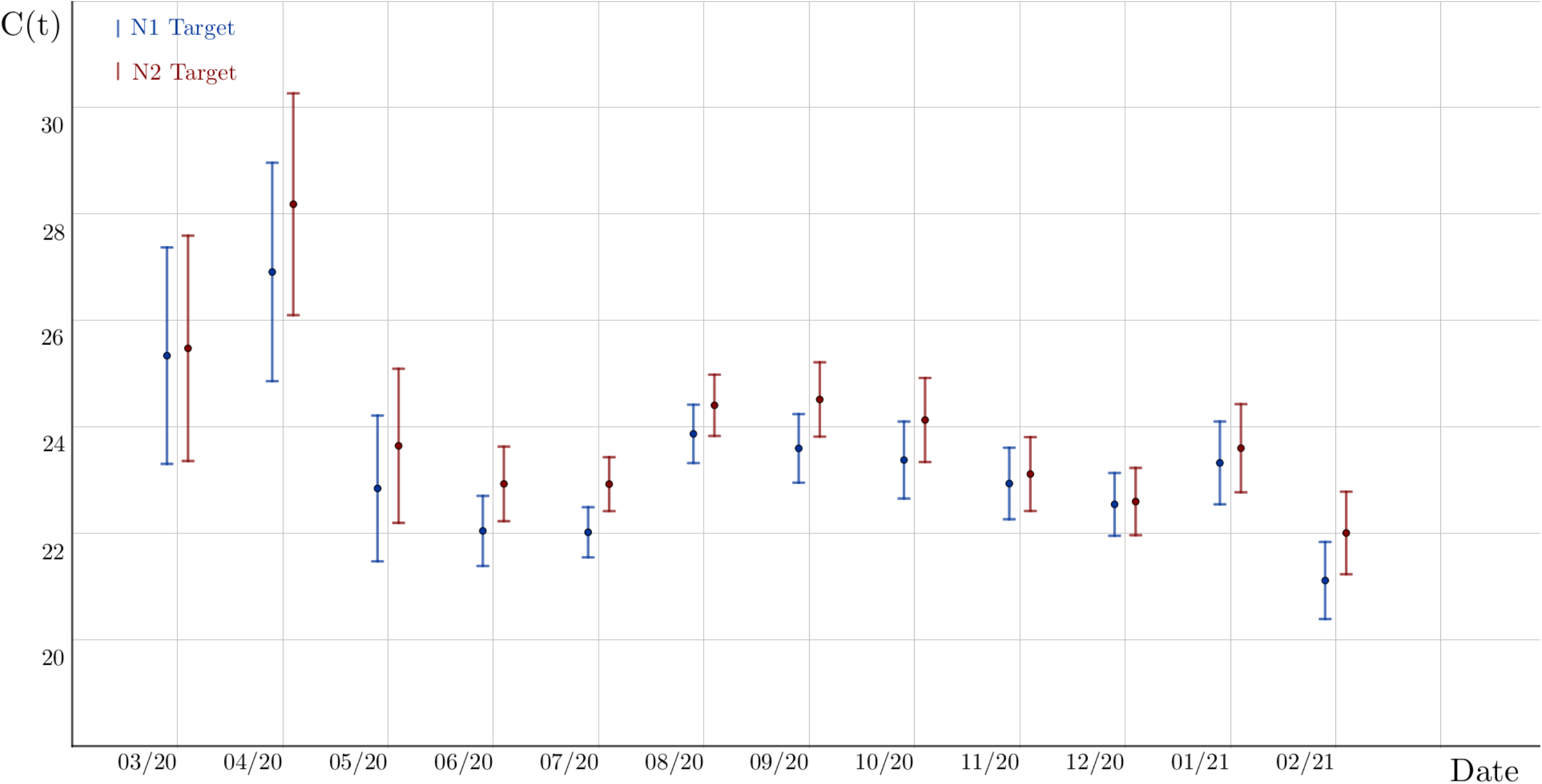
Cycle threshold (Ct) values of N1 and N2 targets in RT-qPCR performed at Hospital de Clínicas de Porto Alegre from March 2020 to February 2021. Ct values of N1 and N2 were lowest in specimens from February (mean, 21.11; 95% Confidence Interval, 20.39- 21.84; P=0.14 and 22.01; 95% Confidence Interval, 21.23-22.78; P<0.001, respectively) when P.1 lineage was detected in 24 (88.9%) of 27 specimens.

Until January 2021, the distribution of SARS-CoV-2 lineages observed in patients treated at our COVID-19 referral center was similar to that observed in regional reports, which tended to follow the distribution of most Brazilian regions, except for the North, where lineage P.1 had been predominant since December [5,8]. In February 2021, however, an overwhelming increase in hospitalizations has been observed since epidemiological week 6, coinciding temporally with the finding that lineage P.1 became predominant, accounting for the vast majority of specimens selected for genotyping in this period.

Since the hospitalization dynamics observed in Rio Grande do Sul resemble those noted in December in the Northern states, where P.1 vastly predominated among tested specimens, we hypothesize that P.1 might be at least partially associated with the striking rise in hospitalization in Rio Grande do Sul. Previous findings suggesting that P.1 may be more transmissible than other lineages corroborate our hypothesis [1,8,9]. Social behavior and the lack of strict COVID- 19 control measures in RS, as in other Brazilian states, might have been the perfect scenario for dissemination of a more transmissible SARS-CoV-2 VOC.

Our data have limitations and must be interpreted accordingly. First, our sample comprises patients from a single referral center, and the specimens were chosen by convenience; thus, they should not be interpreted as a prevalence study for our region. Nonetheless, it should be noted that the criteria for specimen selection of isolates from January (one P.1) and February 2021 (24 P.1) were the same. Second, although consistent with previous findings [1,9], the lower Ct values observed in February must be interpreted with caution, because Ct values were not adjusted for time from onset of symptoms.

In summary, our findings raise concerns regarding a possible association between lineage P.1 and rapid growth in cases and hospitalizations. Despite the relatively low number of specimens analyzed, the finding that the previously undetected P.1 lineage now accounts for almost 90% of specimens from a COVID-19 referral hospital in a state where exponential growth in hospitalizations has been observed warrants close attention and highlights the need for VOC surveillance. Further studies are necessary to investigate the potential association of P.1 dissemination with rapid increase in hospitalizations.

## Data Availability

The sequences were deposited into GISAID database.

https://www.gisaid.org/

## Ethical Statement

This study was approved by the Ethics Committees from Hospital de Clínicas de Porto Alegre (CAAE: 30767420.2.0000.5327).

## Acknowledgements

We thank the staff of “Laboratório de Diagnóstico de SARS-CoV-2” as well as the staff of the Hospital Infection Committee of our institution (“Hospital de Clínicas de Porto Alegre”) for providing data used in this study.

## Authors’ contributions

APZ, PLW, FCZV, FLM, ALB conceived the work design. PLW, FCZV, FLM, CR, FDP performed the NGS experiments. AFM performed bioinformatic analysis and data interpretation. APZ and AKR performed statistical analysis. APZ and AFM wrote the first draft of the manuscript. All authors critically reviewed.

